# Effect of cannabigerol on sleep and quality of life in Veterans: A decentralized, randomized, placebo-controlled trial

**DOI:** 10.1101/2023.08.31.23294611

**Authors:** Chris R Emerson, Courtney E Webster, Eric J Daza, Brett G Klamer, Meghasyam Tummalacherla

## Abstract

**Background/Objective:** This decentralized, randomized, triple-blind, placebo-controlled study evaluated the efficacy and safety of an oral cannabigerol (CBG) formulation in Veterans with sleep issues.

**Methods:** After inclusion, randomization and a two-week run-in phase, participants received CBG (25 mg daily for two weeks, escalated to 50 mg daily for a further two weeks) or placebo. The primary endpoint was change in sleep quality, assessed via the Medical Outcomes Study Sleep Problems Index II (MOS-SS SPI-II). Additional endpoints included change in quality of life (WHODAS-2.0–12), post-traumatic stress disorder symptoms (PCL-5) and sleep actigraphy data. Safety was assessed based on adverse event reporting.

**Results:** A total of 63 participants were randomized to receive CBG (n=33) or placebo (n=30) and formed the intention-to-treat (ITT) population. Of these, 35 completed the study without major protocol deviations (CBG [n=18]; placebo [n=17]) and formed the per-protocol (PP) population. During active treatment (between day 14 and day 42) MOS-SS SPI-II scores declined numerically (indicating improved sleep) in both treatment groups (in both the ITT and the PP populations) with no statistically discernible difference between the CBG and placebo groups. Similar patterns were observed for WHODAS-2.0–12 and PCL-5 scores. Actigraphy data indicated no discernible difference in sleep patterns between the treatment groups. Five mild, nonserious, adverse events were reported with CBG.

**Conclusions:** Both CBG and placebo tended towards sleep and QoL improvement in Veterans. While no firm conclusion on the efficacy of CBG in improving sleep can be made, the favorable safety profile supports future studies to investigate the benefit of CBG.

## Introduction

While one out of three people report trouble sleeping in their lifetime, the Veteran population is particularly vulnerable to sleep-related disorders, being up to six times more impacted by sleep-related issues than the general population.^1^ The United States Veterans Health Administration (VHA) considers sleep issues among Veterans a healthcare crisis. Insomnia diagnoses for Veterans nearly doubled from 2012 to 2018, and sleep-related breathing disorders increased four-fold over the same time period.^2^ Recent surveys of Veteran populations report a high prevalence of disordered sleep, with 11.4% reporting clinical insomnia and subthreshold insomnia in a further 26.0%,^3^ while others report even higher proportions.^4^ There is abundant clinical evidence that sleep influences pain, fatigue, mood, cognition, and daily functioning. Veterans diagnosed with sleep disorders commonly have comorbidities such as obesity, diabetes, congestive heart failure, depression, post-traumatic stress disorder (PTSD) or traumatic brain injury (TBI).^4^ Studies have demonstrated that sleep target intervention in PTSD patients results in significant remission in both PTSD symptoms and sleep disturbances.^5^

Medicinal cannabis is most commonly used to manage pain, anxiety, and sleep problems. Both hemp and cannabis have been used medicinally for thousands of years, though the scientific and clinical literature is obscured by unreliable or anecdotal reports. Cannabis contains hundreds of active compounds.^6^ Delta-9-tetrahydrocannabinol (Δ9-THC) is the primary psychoactive component and makes up almost 95% of cannabis sales,^7^ while non-psychoactive cannabinoids such as cannabidiol (CBD), cannabinol (CBN) and cannabigerol (CBG) are gaining in popularity for both recreational and clinical use. One particular highly purified CBD oil formulation (Epidiolex®) is FDA approved for use in certain pediatric seizure disorders (Lennox-Gastaut syndrome and Dravet syndrome).^8–10^

Growing research indicates that cannabis is used by Veterans with PTSD to cope with sleep disturbances and other PTSD symptoms.^11,12^ Whole plant cannabis and varying ratios of Δ9-THC/CBD have been studied for their effects on sleep.^13–19^ While CBG exhibits similar activity and affinity characteristics as Δ9-THC and CBD on cannabinoid receptors, CBG has uniquely high (nanomolar to sub-nanomolar) affinity as an α-2 adrenoceptor agonist.^20^ α-2 agonists exhibit antihypertensive, sedative, and analgesic activity with broad clinical effects, and are used to manage hypertension, anxiety, attention deficit hyperactivity disorder (ADHD), chronic pain, and to manage withdrawal symptoms from opiates, benzodiazepine, alcohol, cocaine, and tobacco.^21^ Established α-2 agonists such as clonidine show demonstrated clinical benefit for insomnia in children with ADHD, and some efficacy in improving sleep in adults with PTSD.^22^ While such pharmacology provides support for reports of improved sleep in CBG users,^23^ clinical efficacy data are sparse.

The aim of the present clinical study was to determine whether a daily dose of orally self-administered CBG affects sleep quality and quality of life (QoL) in Veterans and to evaluate safety and tolerability. We also investigated the effect of CBG on sleep, activity, and heart rate biometrics. Our goal was to establish feasibility and generate preliminary findings that can be used for larger trials and future research.

## Methods

### Study design and population

This was an interventional, prospective, triple-blinded, randomized and placebo-controlled trial investigating the efficacy, safety, and tolerability of short-term use of a CBG formulation on insomnia and QoL in Veterans ≥21 years of age with self-reported problematic sleep. The study was conducted between October 26, 2021 and May 10, 2022. Study participants were equally randomized, then entered a two-week run-in phase. Participants then received allocated treatment (25mg CBG daily or placebo for two weeks, with escalation of dosing to 50mg daily for the final two weeks) (**Figure 1**). Sleep quality and QoL outcomes were assessed throughout the study via established questionnaires and daily diaries.

**Figure 1.**
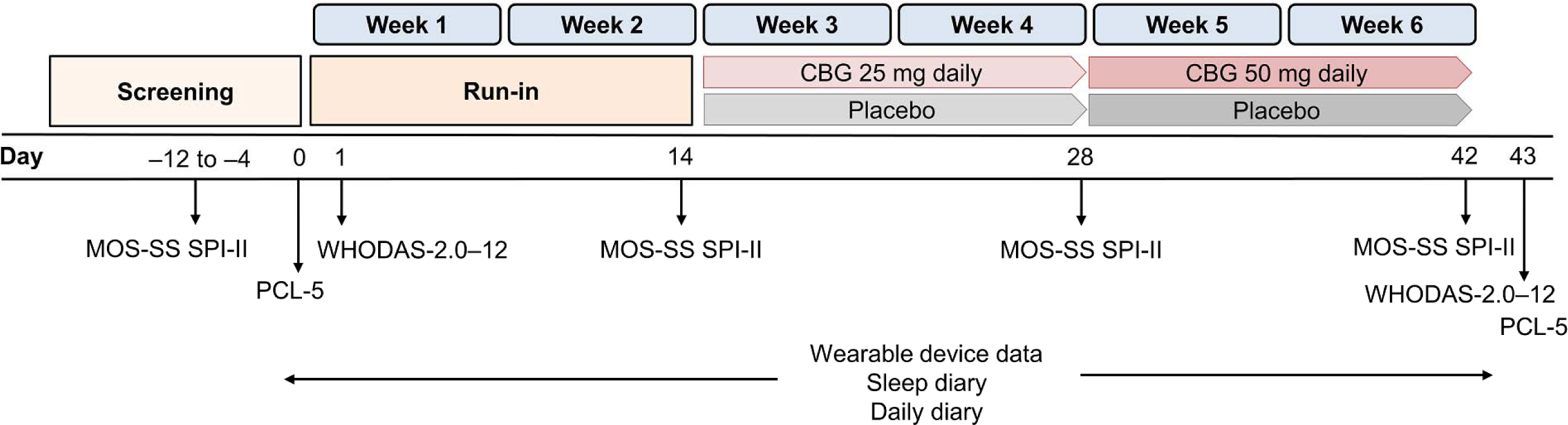
Study design and schedule of assessments. Following screening, all randomized participants entered a two-week run-in phase, then received allocated treatment (CBG 25 mg once daily or placebo for two weeks with escalation of dosing to 50 mg daily for the final two weeks). MOS-SS SPI-II was self-reported at baseline, and on day 14, day 28 and day 42. WHODAS-2.0–12 and PCL-5 were self-reported at baseline (day 0/1 and day 43). Participants maintained daily sleep and evening diaries including allocated medication compliance throughout the study. Fitbit-based outcome measures (sleep and activity duration, heart rate) were monitored from the initial run-in phase to end of study. Abbreviations: CBG, cannabigerol; MOS-SS SPI-II, Medical Outcomes Study Sleep Scale Sleep Problems Index II; PCL-5, PTSD Checklist for the Diagnostic and Statistical Manual of Mental Disorders-Fifth Edition; WHODAS-2.0–12, 12-item version of World Health Organization Disability Assessment Schedule, version 2.0.

Our primary objective was to estimate and evaluate the effect of CBG on the mean change in sleep quality, evaluated using the self-reported Sleep Problems Index II subscale of the Medical Outcomes Study Sleep Scale (MOS-SS SPI-II).^24,25^ Our secondary objective was to evaluate the effect of CBG on QoL, assessed using the short 12-item form of the World Health Organization Disability Assessment Schedule, version 2.0 instrument (WHODAS-2.0–12).^26^ An exploratory objective was to evaluate the effect of CBG on PTSD symptoms, evaluated via the PTSD Checklist for the Diagnostic and Statistical Manual of Mental Disorders-Fifth Edition (PCL-5).^27^ Additional exploratory objectives included evaluation of sleep, activity, and heart rate biometric data, collected via a commercial wrist-worn activity tracking device (Fitbit Inspire 2).^28,29^ For safety evaluation, subjective study product effects, psychological distress and adverse events (AEs) were monitored throughout the study.

This was an entirely decentralized clinical trial (DCT) conducted across California. Participants were recruited across California through Veteran’s associations and advocacy groups (including the Veterans Cannabis Group, Tactical Patients Group, Operation EVAC, the Santa Cruz Veterans Alliance). Campaigns to raise awareness of the study to potential participants was via several routes including email, printed materials, and study advertisements at point-of-sale cannabis product dispensary locations (**Supplementary Figure 1**). In addition, the study design was informed by insight panels to better understand Veterans’ lived experience and potential obstacles to study recruitment and retention. A remote clinical trial management system (Curebase Inc) was used for eligibility assessment, screening, consent, randomization, deployment of study questionnaires, and subsequent data collection.^30^ Each participant was assigned a dedicated clinical research coordinator (CRC).

Inclusion and exclusion criteria are shown in **Supplementary Table 1**. Inclusion criteria included male and female Veterans ≥21 years of age, resident in California, capable of giving informed consent, and who possessed an internet enabled smartphone. Female participants of childbearing potential were required to use an effective form of birth control. Participants were excluded if they were receiving cognitive behavioral therapy (CBT) for insomnia. Participants receiving sleep medications were included if medication and dosage were stable (no medication or dose change in four weeks). Similarly, continuation with existing other psychotropic medications was allowed if medication and dosage were stable. Potential participants with obstructive sleep apnea (OSA) were eligible if the use of continuous positive airway pressure (CPAP) or alternative PAP devices was established for more than four weeks prior to study entry. The presence of PTSD was not an exclusion criterion, although as outlined below PTSD symptom severity was monitored across the study. Use of cannabis products was allowed, with self-reported use assessed at baseline including regularity, frequency of use (daily, multiple times etc.) and product type (Δ9-THC, CBD or both). Following initial eligibility screening, prospective participants were asked to complete the MOS-SS SPI-II survey with a cutoff score of ≥30 used as a proxy indicator of sleep disturbance (broadly comparable, although slightly lower than that used in previous studies in US Veterans, where a threshold of ≥35 was used).^31,32^

The study was conducted in accordance with the Declaration of Helsinki and the International Conference on Harmonization Good Clinical Practice Guidelines (ICH-GCP). The study protocol and supporting documentation was approved by a central Institutional Review Board (Advarra; Pro00056526) and registered at clinicaltrials.gov (NCT05088018). Written informed consent was obtained from each participant prior to enrollment. Study participants received no remuneration or reimbursement, although were allowed to retain their Fitbit device at the end of the study.

### Investigational product

The investigational product was a commercially available CBG formulation (Protab^TM^ by LEVEL) extracted from high CBGa expressing *Cannabis sativa* cultivars prior to senescence. Extraction is via a closed-loop hydrocarbon process entirely preserving the CBG acidic component (CBGa) whilst removing most terpenoids, flavonoids, and other extraneous moieties. This generates a highly enriched CBGa fraction, with further purification and verification steps and analysis for potency conducted through to final formulation. Once formulated, tablets are created on a rotary tablet press, with inline production sampling and analysis to ensure product quality control. Product packaging includes information about the date of production, lot number, batch number, dosage, and storage instructions. For the present study, each batch of the investigational product was evaluated by an independent laboratory to ensure that the cannabinoid profile was within established limits and without Δ9-THC.

Optimal dosing for CBG remains as yet ill-defined; commercially available products have CBG quantities ranging from a few milligrams to up to 33 mg per dose. For the present study we selected a target dosage range that mirrored commonly available dosage recommendations, with an upper range of 50 mg daily, formulated as 25 mg tablets. All participants received two shipments of study product; the first during the run-in phase included two packages, each containing 10 tablets (20 doses total). The second shipment during the initial part of the treatment phase included three packages, each containing 10 tablets (30 doses total). Packaging, labeling and dosage administration of the placebo formulation were identical to that used with the investigational product. All packages can be stably stored at room temperature. After the two-week run-in phase, participants allocated to receive CBG were instructed to take one tablet daily (25 mg) for two weeks (anytime during the day but no more than three hours before bedtime). This caveat was based on empirical reports received by the study principal investigator (C.R.E.) via personal communication with industry colleagues that CBG might increase dream intensity. As such, bedtime dosing was intentionally avoided to reduce any potential adverse impact on dream-state. At the end of this two-week period, participants were then instructed to take two tablets together each day (50 mg) for the following two weeks. Those allocated to placebo followed the same schedule (and similar restricted bedtime use).

### Randomization, Masking, and Data management

Following screening and acceptance into the study, participants were equally randomized across both treatment arms via stratified block randomization, balanced for reported sex (male: female; 9:1) to be consistent with the target Veterans population.

The study was triple-blinded to treatment allocation. Access to randomization lists was restricted to the logistics team responsible for distributing the study product, a Curebase solutions engineer, and the medical monitor. All participants and their assigned CRC were blinded to treatment assignment, as were all study investigators, and the study analyst. All study questionnaire data and reported adverse events were captured via a web-based electronic data capture system for entry into electronic case report forms (eCRFs). This system also provided each participant with a study calendar and reminders for their check-in calls with their assigned CRC and for questionnaire completion and submission.

### Outcome measures and endpoints

Baseline demographics and clinical characteristics were assessed at inclusion (at pre-screening) or at the start of the two-week pre-treatment run-in phase. A complete list of study assessments at different timepoints is shown in **Supplementary Figure 2**).

The primary outcome was the change in participant sleep quality measured using the MOS-SS SPI-II survey.^24,25^ This is a long-established instrument in assessing sleep quality. It was initially validated in a large, heterogeneous population comprising 3,445 adults with chronic conditions that exhibited high incidence rates of insomnia and sleep disturbances, with a mean MOS-SS SPI-II composite score of 29.15 (± standard deviation [SD] 18.04).^24,25^

MOS-SS SPI-II assesses self-reported sleep quality during the preceding two weeks across four domains; sleep disturbance (four items), shortness of breath or headache upon awakening (one item), sleep adequacy (two items), and daytime somnolence (two items), reflecting symptoms consistent with insomnia. Responses are rated on a 6-point Likert scale then transformed and averaged across items to generate a standardized composite score ranging from 0–100. Higher scores indicate greater sleep impairment.^25^ MOS-SS SPI-II has high test-retest reliability with an intraclass correlation coefficient (ICC) of 0.86 (95% confidence interval [CI]. 0.75–0.91) reported in patients with neuropathic pain.^33^

MOS-SS SPI-II has been used to evaluate sleep quality in Veterans including those with post-traumatic stress disorder (PTSD) and traumatic brain injury (TBI). A recent RCT evaluating the impact of holistic yoga or a wellness lifestyle program on insomnia in Veterans exhibiting PTSD reported baseline MOS-SS SPI-II values (± SD) of 57.0 ± 16.8 and 60.0 ± 17.5.^34^ Another study involving Veterans with mild PTSD receiving mind–body interventions or sleep education for insomnia reported baseline mean ± SD values of 56.4 ± 16.6 and 61.4 ± 14.1,^31^ while a similar study conducted in Veterans with mild TBI reported values >60.^32^ There is no generally established minimal clinically important difference (MCID) for MOS-SS SPI-II. In one recent RCT evaluating the impact of holistic yoga or a wellness lifestyle program on insomnia in Veterans exhibiting PTSD baseline MOS-SS SPI-II values (± SD) of 57.0 ± 16.8 and 60.0 ± 17.5 were reported points.^34^ Use of yoga resulted in a 12-point decrease in MOS-SS SPI-II after 16-weeks, while a decrease of 4.6 points was observed in the wellness lifestyle program comparator.^34^

While the conventional MOS-SS SPI-II questionnaire evaluates self-reported sleep quality over the past four weeks (i.e., using a past four-week recall interval),^24,25,35^ modifications to evaluate sleep across shorter periods (e.g., over a one or two-week time frame) are feasible and validated.^31,36^ For the present study, we chose a two-week timeframe. MOS-SS SPI-II responses were collected at baseline (pre-screening), day 14 and after two and four weeks (day 28 and day 42 respectively) (**Figure 1**). The primary endpoint was a “change score” specified as the mean change in MOS-SS SPI-II from the two-week pre-treatment run-in period (day 14) to four weeks after the start of treatment (i.e., day 42, end of study; i.e., six weeks from the baseline).

The secondary outcome was evaluating change in participant QoL measured using WHODAS-2.0 (assessing QoL over the preceding 30 days).^26,37,38^ The conventional WHODAS-2.0 instrument contains 36-items assessing disability and limitations experienced over the preceding 30 days across six domains: cognition, mobility, self-care, getting along with others, life activities, and participation, each rated on a 5-point scale ranging from 0 (No Difficulty) to 4 (Extreme Difficulty/Cannot Do). This instrument shows good reliability and validity and sensitivity to change across a range of physical and mental health conditions,^39,40^ and is increasingly used as an outcome measure in recent years.^37^ including studies in US Veteran populations with or without PTSD.^41,42^ An abbreviated 12-item version (WHODAS-2.0–12) in which each domain has two items is also available,^26^ and used in approximately one-third of studies, ^37^ including studies in US Veterans.^38^

In the present study we chose to use the shorter WHODAS-2.0–12 (using the simple scoring method) in which the summed totals across domains generate a global functional disability score ranging from 0 (no disability) to 48 (complete disability). One study evaluating CBT in patients with stress and anxiety disorders reported baseline WHODAS-2.0–12 scores (using simple scoring) of 23.4 ± 7.9.^43^

This secondary endpoint was also a change score, specified as the mean change in WHODAS-2.0–12 from the run-in period to end-of-study.

We also evaluated PTSD symptoms (via PCL-5) at baseline and at end of study to explore any differences in the effect of CBG on PTSD severity. PCL-5 is a 20-item self-reported inventory of PTSD symptoms over the past 30 days, rated on a 5-point Likert-scale (from 0 = not at all to 4 = extremely) to generate a total scored on a scale of 0–80, with higher scores indicating greater PTSD symptoms.^27^

Participants also completed a sleep diary each morning throughout the study, recording information on how they slept, sleep interruptions, and medication, drug or alcohol use prior to sleeping. An additional daily evening diary was used, asking participants to assess their satisfaction with and productivity of their day, and also to record their adherence to their allocated treatment (number of tablets and time taken).

Exploratory outcomes examined included data on sleep quality (sleep efficiency; time spent in light, deep, and rapid eye movement [REM] sleep stages), activity tracking (intraday steps; time spent in ‘fairly active’, ‘lightly active’, ‘sedentary’, and ‘very active’ activity stages) and resting and active heart rate measures. Data was collected via the Fitbit Inspire 2 device,^28^ where granular ‘intraday’ data collection (to the minute or second level for activity and heart rate) is feasible and was collected.^29^ Following a one-time set up, linking the device to the allied Fitbit smartphone app (with authorization to share data with the study application), periodic data collection from the device is synced with the app via Bluetooth. Participants were instructed to wear the Fitbit at all times (with a recommendation that the device be charged when showering). The resultant passively-collected sleep data allows comparison with each participant’s reported sleep quality.

### Safety and tolerability assessment

Safety monitoring responsibilities were assigned to an independent clinician (a psychiatrist with experience treating persons who have experienced trauma). AEs could be reported at any time through the study application. Participants would be prompted to enter the date of onset, whether the experience was still ongoing, and a brief description. The submission of an AE through the study application would immediately alert the study coordinator team, who would reach out to the participant and gather additional details. The AE would be reviewed by the independent clinician to assess causality and recommend if the participant should continue the study.

The study application was also designed to trigger an alert to an on-call clinician when specific responses in the PCL-5 might indicate the participant is experiencing a current and severe crisis. If a participant responded with “Extremely” to either Question 9 (addressing negative beliefs) “*In the past month, how much were you bothered by: Having strong negative beliefs about yourself, other people, or the world (for example, having thoughts such as: I am bad, there is something seriously wrong with me, no one can be trusted, the world is completely dangerous?)*“ or Question 16 (addressing taking risks/self-harm) “*In the past month, how much were you bothered by: Taking too many risks or doing things that could cause you harm?*”, this would trigger a text message and email alert to the independent clinician. Upon receiving such alerts, the clinician would immediately call the participant, assess the situation, and then recommend sources of emergency or non-emergency support if deemed necessary.

### Sample size

No previous study has reported changes in the primary outcome measure (MOS-SS SPI-II) following any cannabinoid therapy. There is no generally established minimal clinically important difference (MCID) for either MOS-SS SPI-II or WHODAS-2.0–12. In one recent RCT evaluating the impact of holistic yoga or a wellness lifestyle program on insomnia in Veterans exhibiting PTSD, baseline MOS-SS SPI-II values (± SD) of 57.0 ± 16.8 and 60.0 ± 17.5 points were reported.^34^ Use of yoga resulted in a 12-point decrease in MOS-SS SPI-II after 16-weeks, while a decrease of 4.6 points was observed in the wellness lifestyle program comparator.^34^ Similarly, at present, a single MCID indicating a clinically meaningful change in WHODAS-2.0 remains to be established,^37^ although some studies have estimated MCIDs for WHODAS-2.0–12 ranging from 3 to 7 points.^43,44^

Based on this adjacent literature and scientific judgment, sample size was calculated by assuming that a mean 10-point improvement in either the MOS-SS SPI-II or WHODAS-2.0–12 between the two treatment groups would be considered as clinically meaningful. In addition, the largest SD of pre- and post-treatment outcomes (not endpoints) for either outcome in either treatment group was assumed to be 20.95. From this, it was estimated that an overall initial sample size of 50 participants per arm (accounting for 40% attrition before study completion) would provide ≥80% power, with an alpha level (i.e., maximum false positive rate, statistical significance level) of 0.05 for a two-sample, two-sided t-test.

### Data presentation and analysis

Descriptive analyses were performed for all study variables. Continuous variables are presented as mean values (± SD) or median and interquartile range (IQR) i.e., first quartile to third quartile (Q1–Q3). Categorical variables are presented as frequency counts and percentages. Analyses were performed for both the intention-to-treat (ITT) and the per-protocol (PP) populations. The ITT population included all participants who were randomized at baseline regardless of subsequent compliance or withdrawal. The PP population included all participants without any major protocol deviations (i.e., study withdrawal or loss to follow-up, failure to complete study questionnaires, and non-compliance with treatment administration schedule). Safety analyses were conducted in the entire population by tabulating adverse events (i.e., statistical description and summary only, no inference or testing).

Differences in baseline characteristics between the treatment groups were assessed using either Pearson’s Chi-squared test, Fisher’s exact test, or the Wilcoxon rank sum test. Mean values at day 14 and after four weeks (day 42) for both treatment groups were computed. The endpoint was estimated for each group by calculating the mean change score across all participants. The treatment effect for this endpoint was then estimated as the difference in the mean endpoint for the CBG group minus the mean endpoint for the placebo group, evaluated using Welch’s t-test. A similar approach was used for the secondary endpoint evaluating WHODAS-2.0–12 change from the run-in period to end-of-study (comparing differences between day 43 and day 1), and for our exploratory PCL-5 evaluations (comparing differences between day 43 and baseline [day 0]).

Planned exploratory analyses included characterizing the effect of different CBG dosing on change in MOS-SS SPI-II, evaluated by assessing the initial early effect after two weeks of 25 mg daily treatment (between day 14 and day 28) and comparing this with the overall change after all four weeks of treatment (i.e., with 25 mg CBG daily for two weeks and then 50 mg CBG daily for two weeks more). Analyses of Fitbit-based outcome measures (daily averaged sleep and activity duration, heart rate) were assessed using linear mixed-effects models. Each model included subject-level random intercepts and subject-level random slopes over time, along with fixed effects of age, BMI, study day, treatment group, and the baseline outcome value. All statistical analyses were conducted in R version 4.2.2 (R Core Team).

### Post-hoc analysis

We conducted post-hoc analyses to evaluate characteristics of participants receiving CBG responding to treatment, defined as those individuals with a 10-point improvement (decrease) in their MOS-SS SPI-II score between day 14 and day 42.

## Results

### Study population and participant disposition

A total of 407 individuals were assessed for eligibility, of which 205 were excluded due to failure to meet eligibility criteria and a further 139 declined to participate (**Figure 2)**. Sixty-three participants were randomized to allocated dosing; CBG (n=33) and placebo (n=30) and comprised the ITT population. Of these, 8 participants (12.7%) failed to complete the study for the following reasons; withdrew for personal reasons or lost to follow up (n=3); withdrew due to AEs (n=2) and lack of compliance or dosing adherence (n=3). The PP population comprised a total of 33 participants; CBG (n=18) and placebo (n=17). Excluded from this analysis set were 20 due to insufficient survey data concerning the primary and secondary endpoints or major dosing protocol violations.

**Figure 2.**
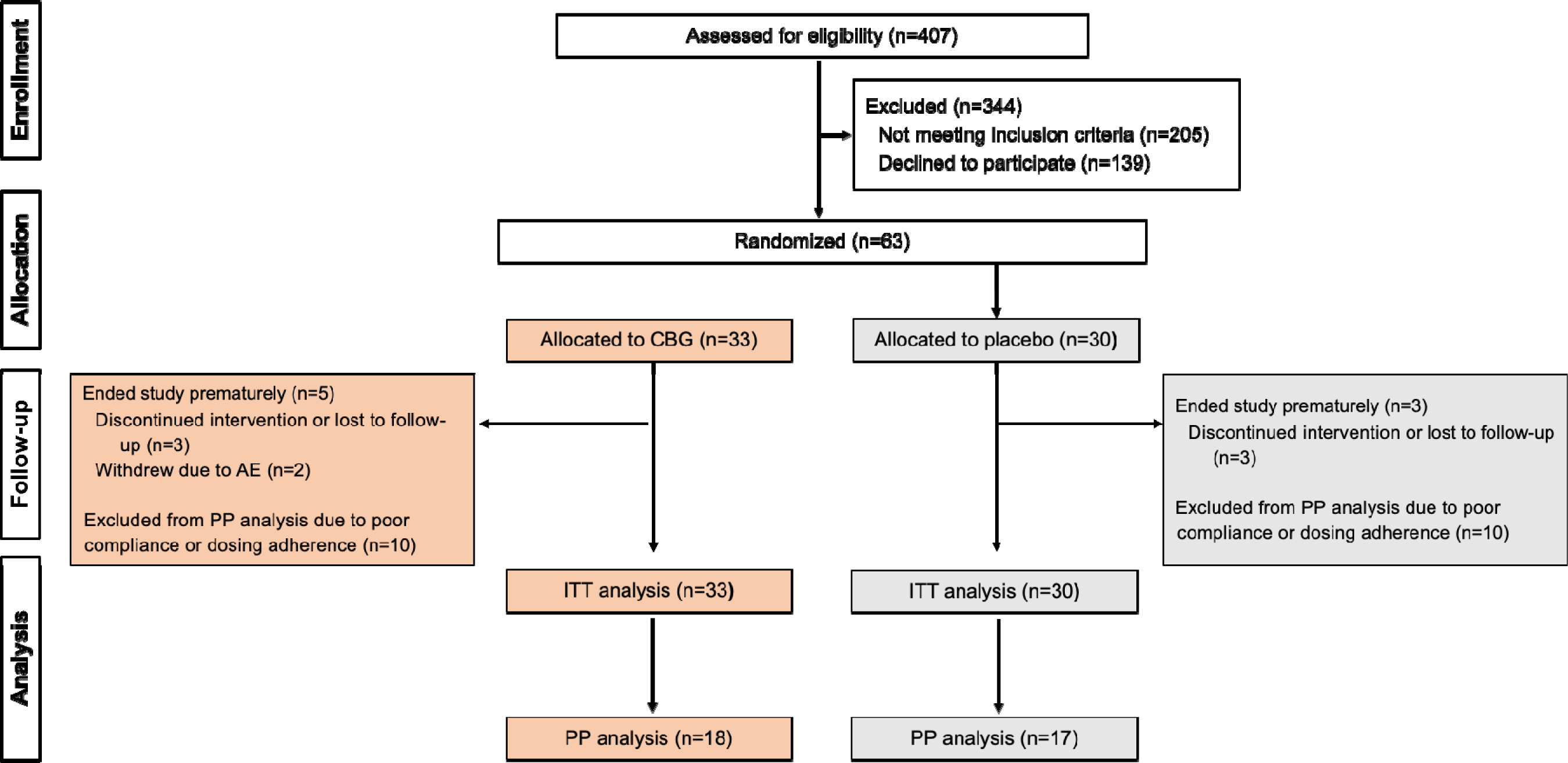
Participant flowchart. Abbreviations: AE, adverse event; CBG, cannabigerol; ITT, intention-to-treat; PP, per-protocol

Baseline demographics and clinical characteristics were comparable across both treatment groups in both the ITT and PP populations (**Table 1**). Participants in the overall study population had a mean age of 44.0 ± 13.6 years (range 23.4–74.1); males accounted for 77.7% and the majority of participants (68.3%) were Caucasian. Most participants had longstanding sleep problems, with 70% reporting problems for more than 5 years. On average, participants reported 5.3 ± 1.3 hours of sleep each night (range 3.0–10.0), with a mean MOS-SS SPI-II at baseline of 61.4 ± 15.4 (range 31.7–91.7). Mean PCL-5 scores were high in both treatment arms although with substantial variation; CBG (32.1 ± 18.9, range 6.0–75.0) and placebo (33.2 ± 19.1, range 1.0–71.0); and over 40% of participants in both groups had scores ≥33 indicating PSTD.^45^ Nine participants (14.3%) reported physician diagnosed OSA, all of whom used CPAP. While regular tobacco use was low (15.9%), the majority of participants (85.7%) reported regular use of one or more cannabis products, with 75% reporting daily use (and often multiple times daily). In this, while around one-third of patients reported use of Δ9-THC only, almost 50% reported use of both Δ9-THC and CBD.

**Table 1.**
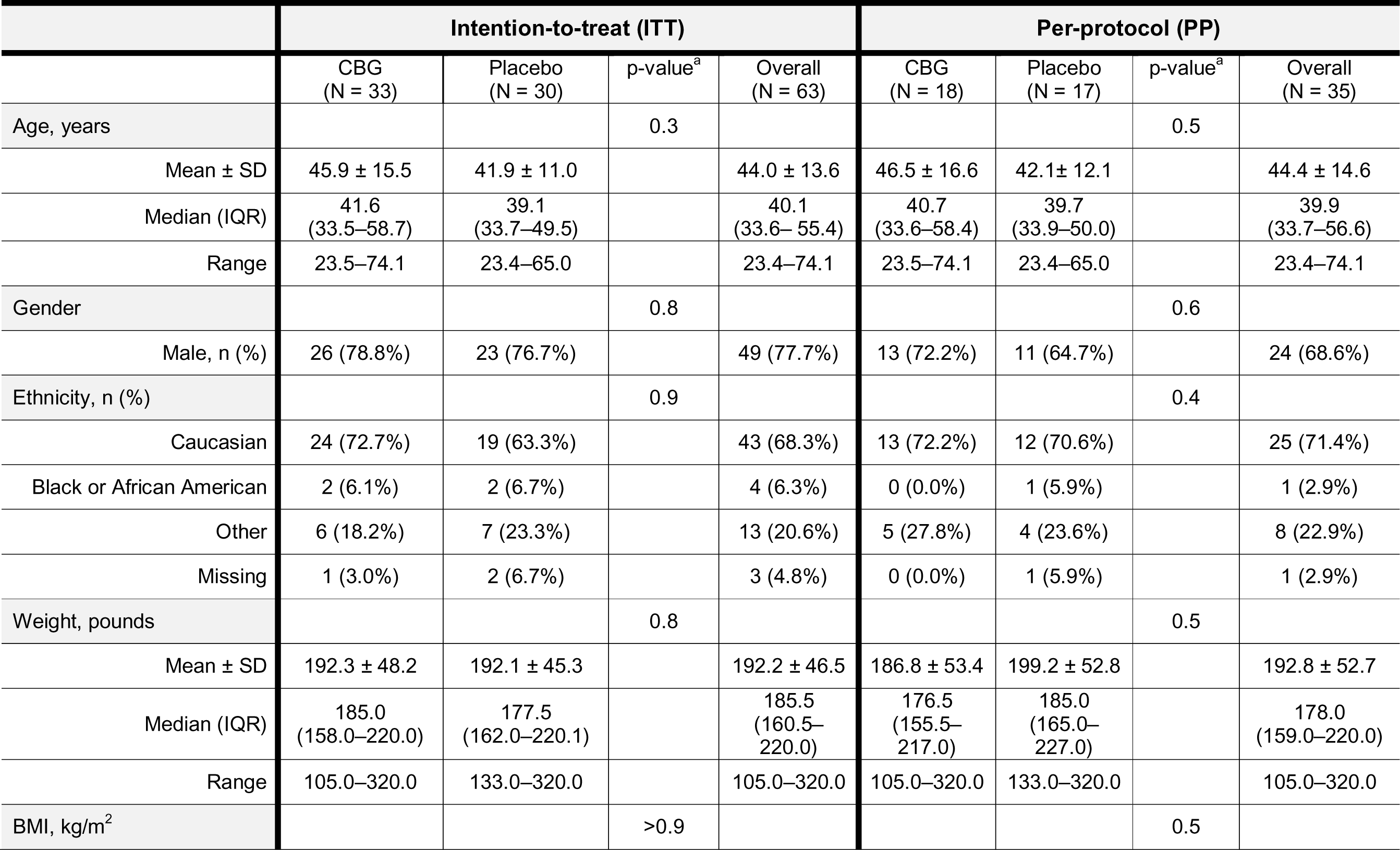
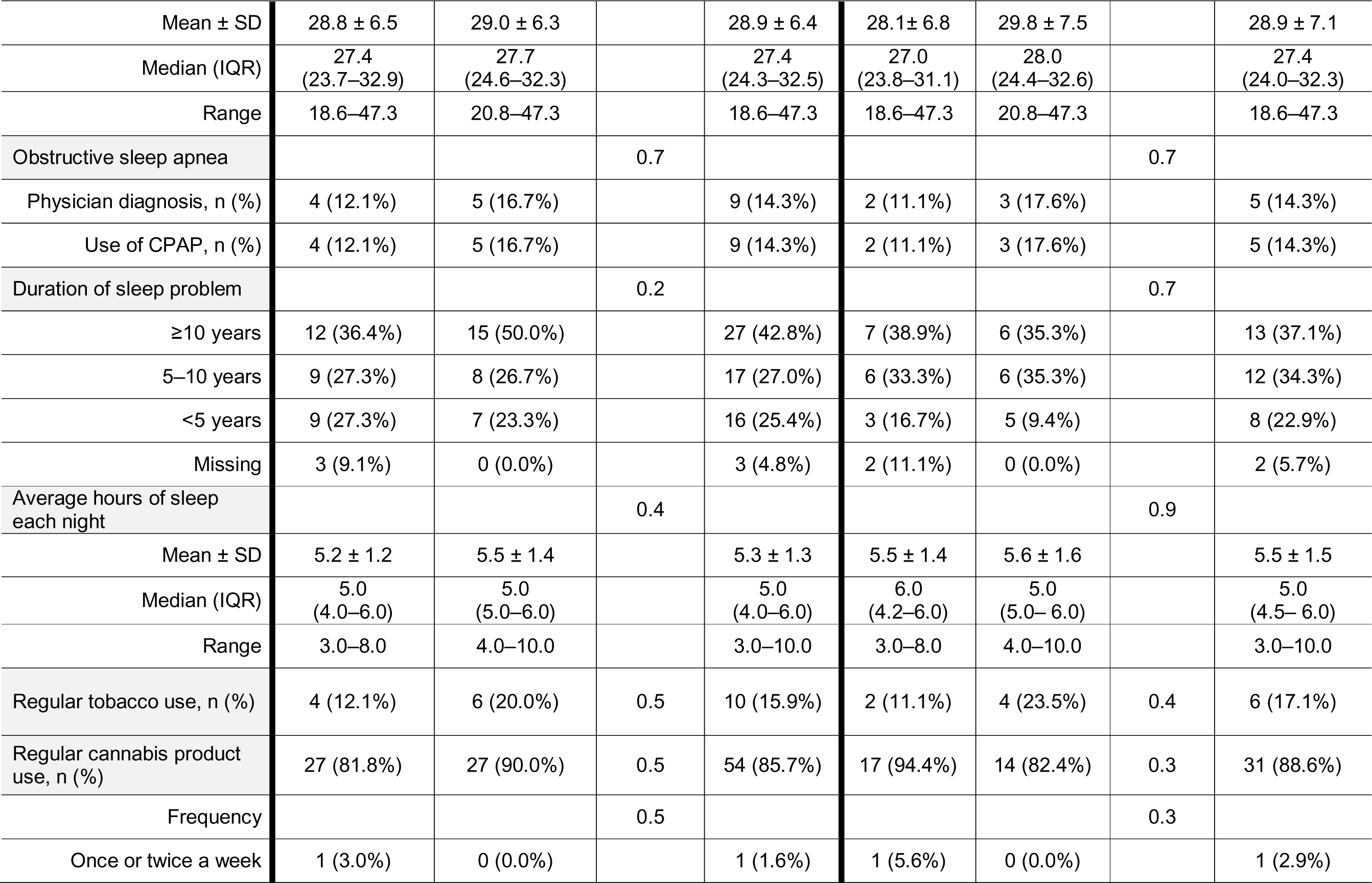
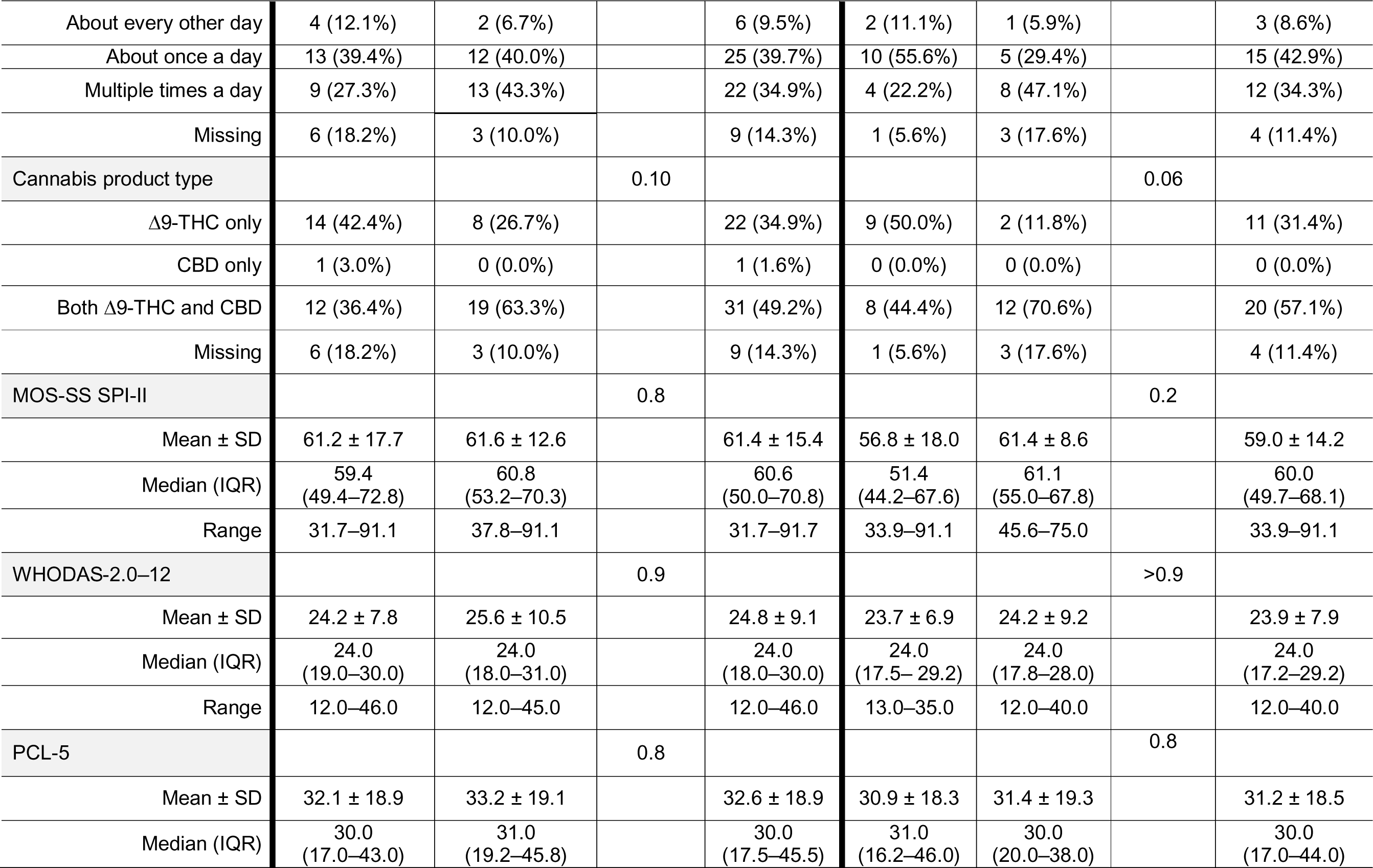
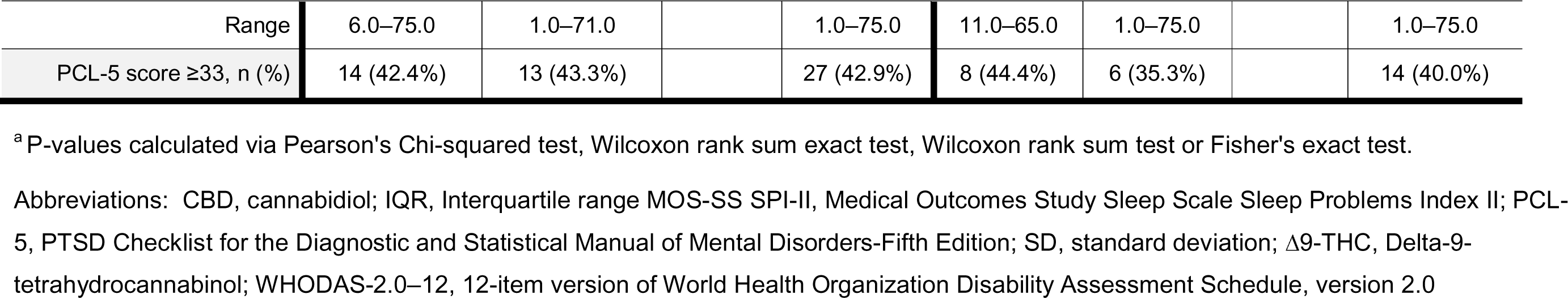
Baseline demographics and clinical characteristics of the study cohort.

### Effect on sleep

MOS-SS SPI-II scores numerically declined (indicating improved sleep) in both treatment groups between day 14 and day 42, evident in both the ITT and PP populations (**Table 2** and **Figure 3**). For the ITT population, the mean change ± SD in MOS-SS SPI-II was a decrease of 7.0 ± 17.1 points (range, –25.0, 58.9) in those participants receiving CBG, and by 11.4 ± 13.0 points (range, –15.6, 54.4) in those receiving placebo. For the primary endpoint, the mean difference of within-group change between groups was 4.4 (95% CI, –4.1 to 13.0) in favor of placebo (*p* = 0.3). Similar results were seen in the PP population, with a mean reduction in MOS-SS SPI-II scores of 9.0 ± 15.6 points (range, –4.5, 58.9) in the CBG group, and 12.8 ± 8.4 points (range, 0.0, 28.9) in the placebo group. For the primary endpoint, the mean difference between groups was 3.8 (95% CI, –4.8 to 12.0) in favor of placebo (*p* = 0.4) (**Table 2** and **Figure 3**). There were no substantial differences in changes in MOS-SS SPI-II scores observed after the first two weeks of treatment (i.e., from day 14 to day 28) compared with the change after all four weeks of treatment in either the ITT or the PP population (**Table 2**).

**Figure 3.**
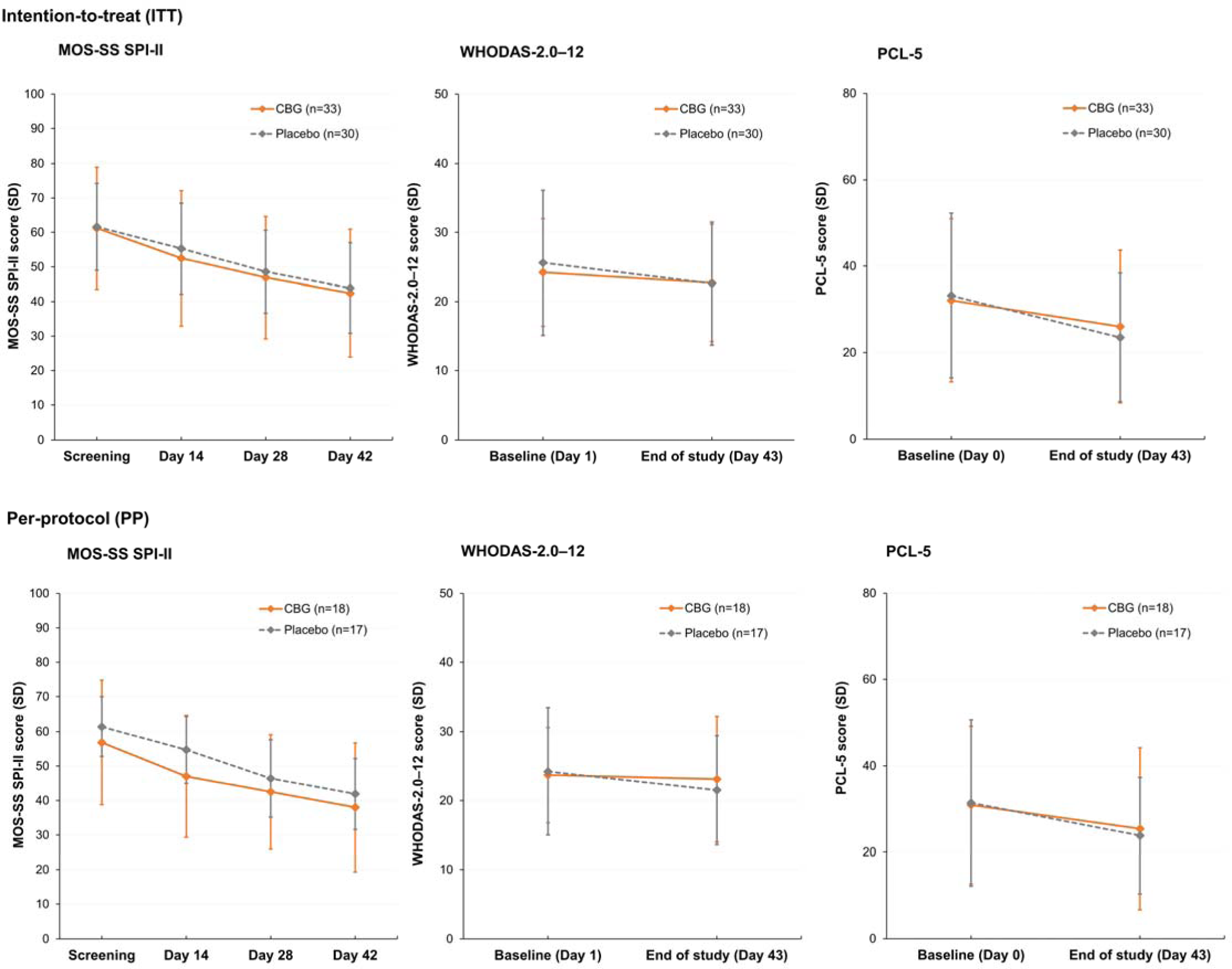
MOS-SS SPI-II, WHODAS-2.0–12 and PCL-5 scores throughout the study. MOS-SS SPI-II was self-reported at screening and on day 14, day 28 and day 42; WHODAS-2.0–12 was self-reported at day 1 and day 43; and the PCL-5 was self-reported at day 0 and day 43. Following screening, all randomized participants entered a two-week run-in phase (through day 14). Participants then received CBG 25 mg daily or placebo for two weeks (from day 15 to day 28) with escalation of CBG dosing to 50 mg daily (from day 29 through day 42). Abbreviations: MOS-SS SPI-II, Medical Outcomes Study Sleep Scale Sleep Problems Index II; PCL-5, PTSD Checklist for the Diagnostic and Statistical Manual of Mental Disorders-Fifth Edition; SD, standard deviation; WHODAS-2.0–12, 12-item version of World Health Organization Disability Assessment Schedule, version 2.0.

**Table 2.**
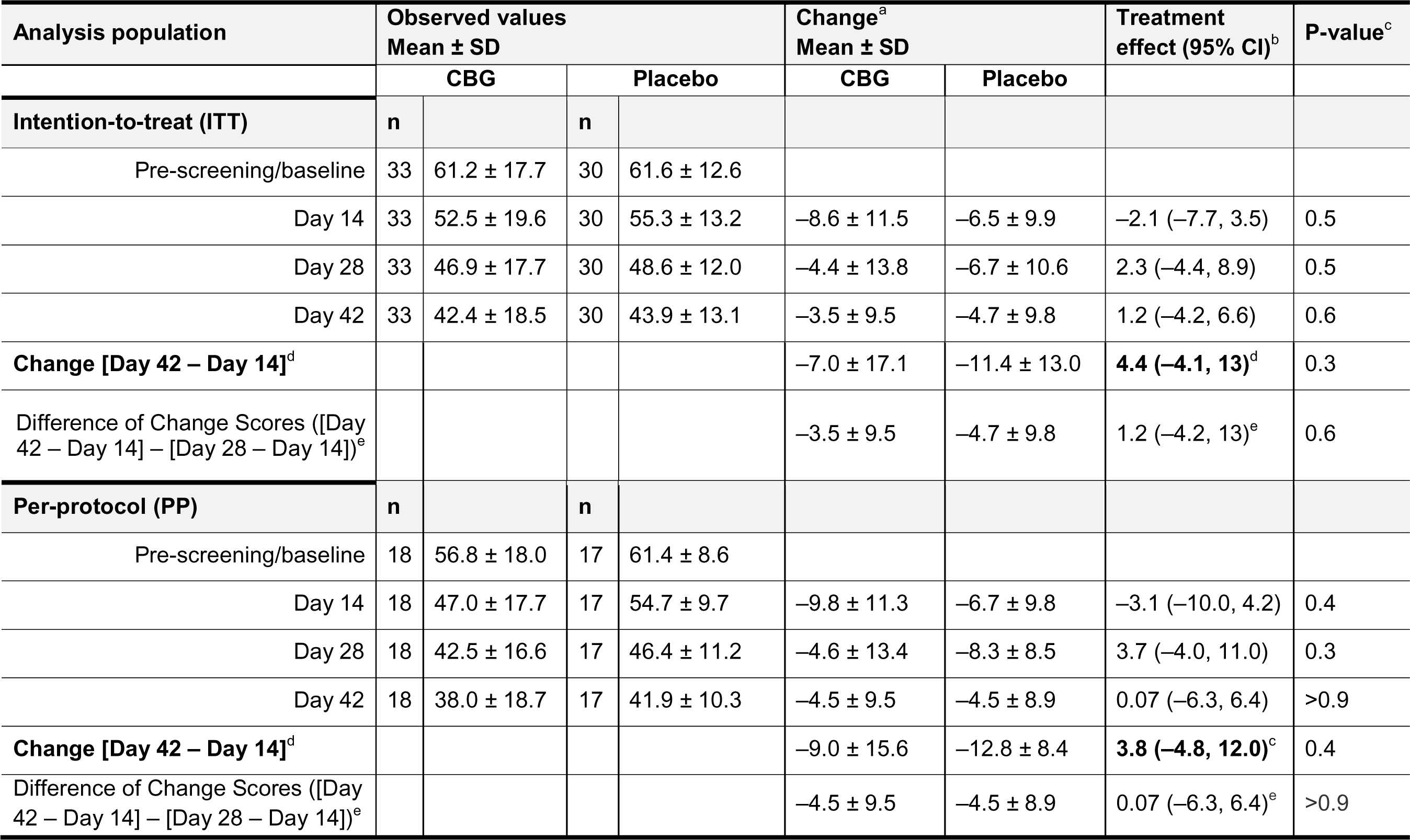
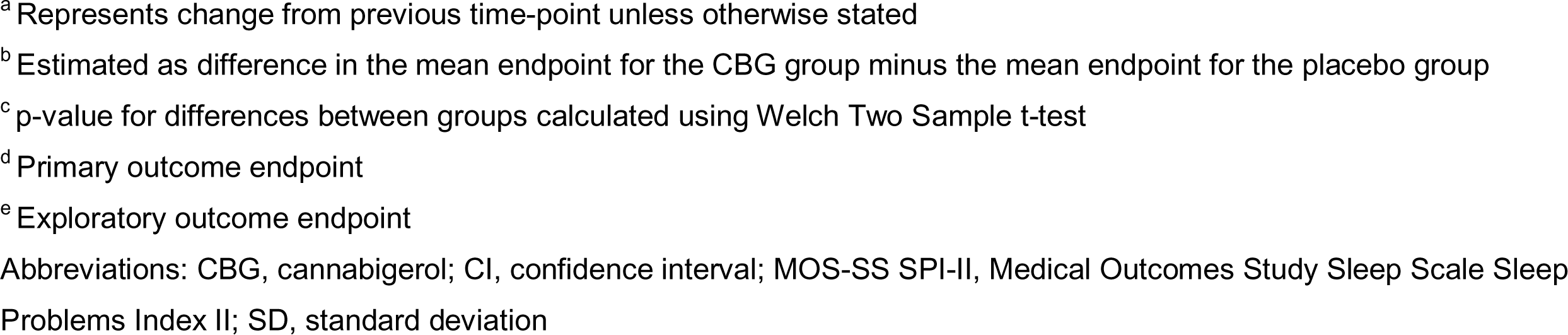
Change in MOS-SS SPI-II scores across the study period.

| Analysis population | Observed values<br>Mean ± SD |  |  |  | Change<sup>a</sup><br>Mean ± SD |  | Treatment<br>effect (95% CI)<sup>b</sup> | P-value<sup>c</sup> |
| --- | --- | --- | --- | --- | --- | --- | --- | --- |
|  | CBG |  | Placebo |  | CBG | Placebo |  |  |
| Intention-to-treat (ITT) | n |  | n |  |  |  |  |  |
| Pre-screening/baseline | 33 | 61.2 ± 17.7 | 30 | 61.6 ± 12.6 |  |  |  |  |
| Day 14 | 33 | 52.5 ± 19.6 | 30 | 55.3 ± 13.2 | −8.6 ± 11.5 | −6.5 ± 9.9 | −2.1 (−7.7, 3.5) | 0.5 |
| Day 28 | 33 | 46.9 ± 17.7 | 30 | 48.6 ± 12.0 | −4.4 ± 13.8 | −6.7 ± 10.6 | 2.3 (−4.4, 8.9) | 0.5 |
| Day 42 | 33 | 42.4 ± 18.5 | 30 | 43.9 ± 13.1 | −3.5 ± 9.5 | −4.7 ± 9.8 | 1.2 (−4.2, 6.6) | 0.6 |
| **Change [Day 42 – Day 14]<sup>d</sup>** |  |  |  |  | −7.0 ± 17.1 | −11.4 ± 13.0 | **4.4 (−4.1, 13)<sup>d</sup>** | 0.3 |
| Difference of Change Scores ([Day 42 – Day 14] – [Day 28 – Day 14])<sup>e</sup> |  |  |  |  | −3.5 ± 9.5 | −4.7 ± 9.8 | 1.2 (−4.2, 13)<sup>e</sup> | 0.6 |
| Per-protocol (PP) | n |  | n |  |  |  |  |  |
| Pre-screening/baseline | 18 | 56.8 ± 18.0 | 17 | 61.4 ± 8.6 |  |  |  |  |
| Day 14 | 18 | 47.0 ± 17.7 | 17 | 54.7 ± 9.7 | −9.8 ± 11.3 | −6.7 ± 9.8 | −3.1 (−10.0, 4.2) | 0.4 |
| Day 28 | 18 | 42.5 ± 16.6 | 17 | 46.4 ± 11.2 | −4.6 ± 13.4 | −8.3 ± 8.5 | 3.7 (−4.0, 11.0) | 0.3 |
| Day 42 | 18 | 38.0 ± 18.7 | 17 | 41.9 ± 10.3 | −4.5 ± 9.5 | −4.5 ± 8.9 | 0.07 (−6.3, 6.4) | >0.9 |
| **Change [Day 42 – Day 14]<sup>d</sup>** |  |  |  |  | −9.0 ± 15.6 | −12.8 ± 8.4 | **3.8 (−4.8, 12.0)<sup>c</sup>** | 0.4 |
| Difference of Change Scores ([Day 42 – Day 14] – [Day 28 – Day 14])<sup>e</sup> |  |  |  |  | −4.5 ± 9.5 | −4.5 ± 8.9 | 0.07 (−6.3, 6.4)<sup>e</sup> | >0.9 |

<sup>a</sup> Represents change from previous time-point unless otherwise stated
<sup>b</sup> Estimated as difference in the mean endpoint for the CBG group minus the mean endpoint for the placebo group
<sup>c</sup> p-value for differences between groups calculated using Welch Two Sample t-test
<sup>d</sup> Primary outcome endpoint
<sup>e</sup> Exploratory outcome endpoint
Abbreviations: CBG, cannabigerol; CI, confidence interval; MOS-SS SPI-II, Medical Outcomes Study Sleep Scale Sleep Problems Index II; SD, standard deviation

Post-hoc analysis was performed for participants receiving CBG (in the ITT population) considered having a clinically meaningful response (in terms of observed decline in MOS-SS SPI-II scores of at least 10 points) during active treatment (i.e., from day 14 to 42) (**Figure 4)**. “Responders” (n=8) were younger (average age 35.2 years ± 11.8) compared to “non-responders” (n=17) with an average age of 50.8 years ± 16.2, (*p* = 0.011), with otherwise non-substantial differences in height, weight, BMI, biological sex, and other characteristics (cannabis and tobacco usage, sleep apnea diagnosis). Furthermore, “responders” did not show meaningful changes in sleep, activity, or heart rate biometrics as reported by Fitbit. Due to the small sample size, we are noting these findings to inform future work.

**Figure 4.**
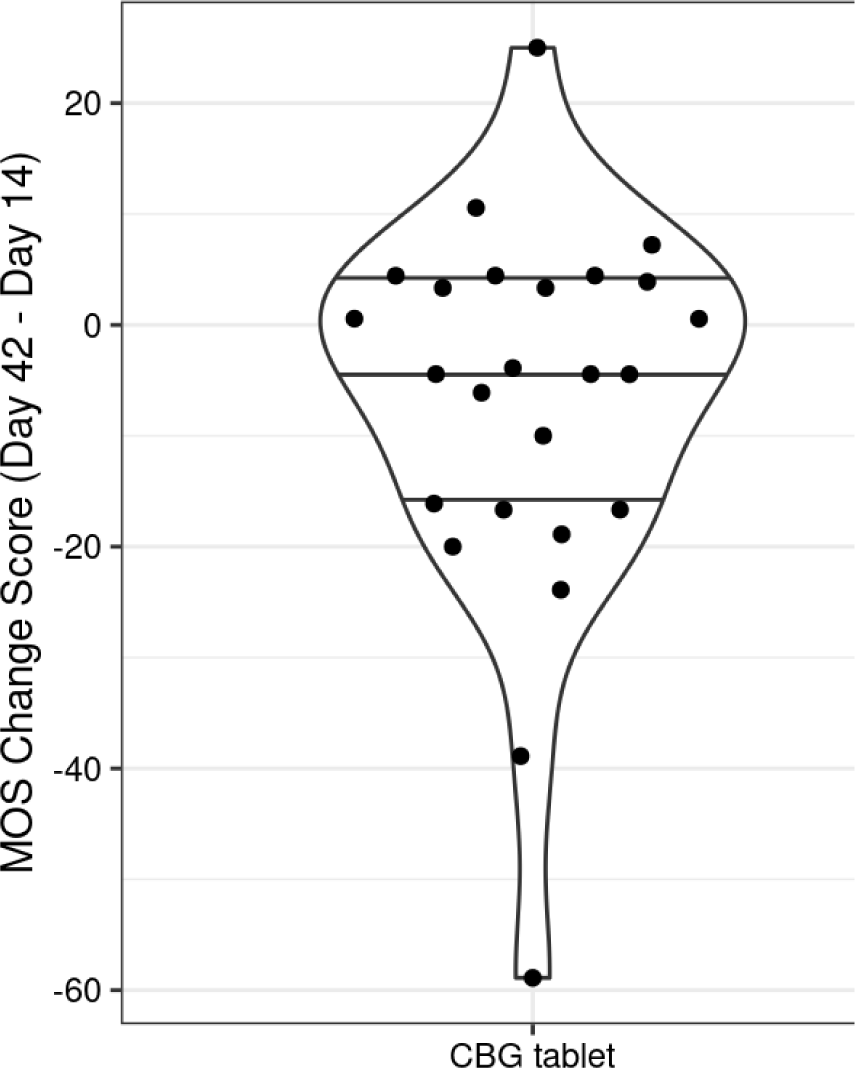
MOS-SS SPI-II responder profile (intention-to-treat population) Funnel plots indicating change in MOS-SS SPI-II during active treatment in participants receiving CBG. Responders were considered those showing an improvement in MOS-SS SPI-II of at least 10-points (considered as clinically meaningful); observed in eight participants receiving CBG. Abbreviations: CBG, cannabigerol; MOS-SS SPI-II, Medical Outcomes Study Sleep Scale Sleep Problems Index II

### Effect on QoL and PTSD symptoms

WHODAS-2.0–12 scores declined in both groups across the study period (indicating improved QoL) although such improvements were small (**Table 3** and **Figure 3**). In the ITT population, WHODAS-2.0–12 scores decreased by 0.8 ± 5.8 points (range, – 12.0, 14.0) in those participants receiving CBG, and by 2.7 ± 4.5 points (range, –4.0, 16.0) in those receiving placebo. The mean difference of within-group change between groups was 1.9 (95% CI, –1.1 to 5.5) in favor of placebo (*p* = 0.2). A similar pattern was observed in the PP population, with a mean group difference of 2.2 (95% CI, –0.83 to 5.3) in favor of placebo (*p* = 0.15) (**Table 3**).

**Table 3.** Change in WHODAS-2.0–12 and PCL-5 scores across the study period.

| Measure/Analysis population | Observed values<br>Mean ± SD |  |  |  | Change<br>Mean ± SD |  | Treatment<br>effect (95% CI)<sup>a</sup> | P-value<sup>b</sup> |
| --- | --- | --- | --- | --- | --- | --- | --- | --- |
|  | CBG |  | Placebo |  | CBG | Placebo |  |  |
| **WHODAS-2.0–12** |  |  |  |  |  |  |  |  |
| **Intention-to-treat (ITT)** | n |  | n |  |  |  |  |  |
| Day 1 | 33 | 24.2 ± 7.8 | 30 | 25.6 ± 10.5 |  |  |  |  |
| Day 43 | 33 | 22.7 ± 8.5 | 30 | 22.6 ± 8.9 |  |  |  |  |
| Change [Day 43 – Day 1]<sup>c</sup> |  |  |  |  | –0.8 ± 5.8 | –2.7 ± 4.5 | 1.9 (–1.1, 4.9)<sup>c</sup> | 0.2 |
| **Per-protocol (PP)** | n |  | n |  |  |  |  |  |
| Day 1 | 18 | 23.7 ± 6.9 | 17 | 24.2 ± 9.2 |  |  |  |  |
| Day 43 | 18 | 23.1 ± 9.1 | 17 | 21.5 ± 7.9 |  |  |  |  |
| Change [Day 43 – Day 1]<sup>c</sup> |  |  |  |  | –0.7 ± 5.0 | –2.9 ± 3.6 | 2.2 (–0.83, 5.3)<sup>c</sup> | 0.15 |
| **PCL-5** |  |  |  |  |  |  |  |  |
| **Intention-to-treat (ITT)** | n |  | n |  |  |  |  |  |
| Day 0 | 33 | 32.1 ± 18.9 | 30 | 33.2 ± 19.1 |  |  |  |  |
| Day 43 | 33 | 26.0 ± 17.7 | 30 | 23.5 ± 14.9 |  |  |  |  |
| Change [Day 43 – Day 1]<sup>d</sup> |  |  |  |  | –5.8 ± 10.8 | –8.7 ± 13.4 | 2.9 (–3.9, 9.7)<sup>d</sup> | 0.4 |
| **Per-protocol (PP)** |  |  |  |  |  |  |  |  |
| Day 0 | 18 | 30.9 ± 18.3 | 17 | 31.4 ± 19.3 |  |  |  |  |
| Day 43 | 18 | 25.4 ± 18.8 | 17 | 23.8 ± 13.5 |  |  |  |  |
| Change [Day 43 – Day 0]<sup>d</sup> |  |  |  |  | -5.5 ± 10.5 | -7.6 ± 13.0 | 2.1 (-6.0, 10.0)<sup>d</sup> | 0.6 |

<sup>a</sup> Estimated as difference in the mean endpoint for the CBG group minus the mean endpoint for the placebo group
<sup>b</sup> p-value for differences between groups calculated using Welch Two Sample t-test
<sup>c</sup> Secondary outcome endpoint
<sup>d</sup> Exploratory outcome endpoint
Abbreviations: CBG, cannabigerol; CI, confidence interval; PCL-5, PTSD Checklist for the Diagnostic and Statistical Manual of Mental Disorders-Fifth Edition; SD, standard deviation; WHODAS-2.0–12, 12-item version of World Health Organization Disability Assessment Schedule, version 2.0.

Improvements in PTSD symptoms, as measured by declines in PCL-5 scores, were also observed in both groups, with greater reductions apparent in the placebo group in both the ITT and the PP populations (**Table 3** and **Figure 3**).

### Effect on activity tracking measures

It is interesting to note that while MOS-SS SPI-II scores improved (decreased) in both groups through the course of the study, wrist actigraphy data indicated that sleep duration, efficiency, minutes asleep, minutes awake, minutes of REM, and overall time in bed were relatively constant (**Figure 5).** While the rationale behind this difference is out of scope in this discussion, biometric monitoring or other real-time, real-world evidence is of continued interest for future studies. One finding of interest was a potential physiological effect of CBG dose timing on resting heart rate. For all time periods except late-night dosing, participants in the ITT population receiving CBG had a lower mean heart rate (2 hours post-dosing) than those taking placebo. In those reporting afternoon dosing (noon–5 PM) a statistically-discernible lower mean heart rate (*p* = 0.017) was observed in the baseline-adjusted model (**Table 4** and **Figure 6**).

**Figure 5.**
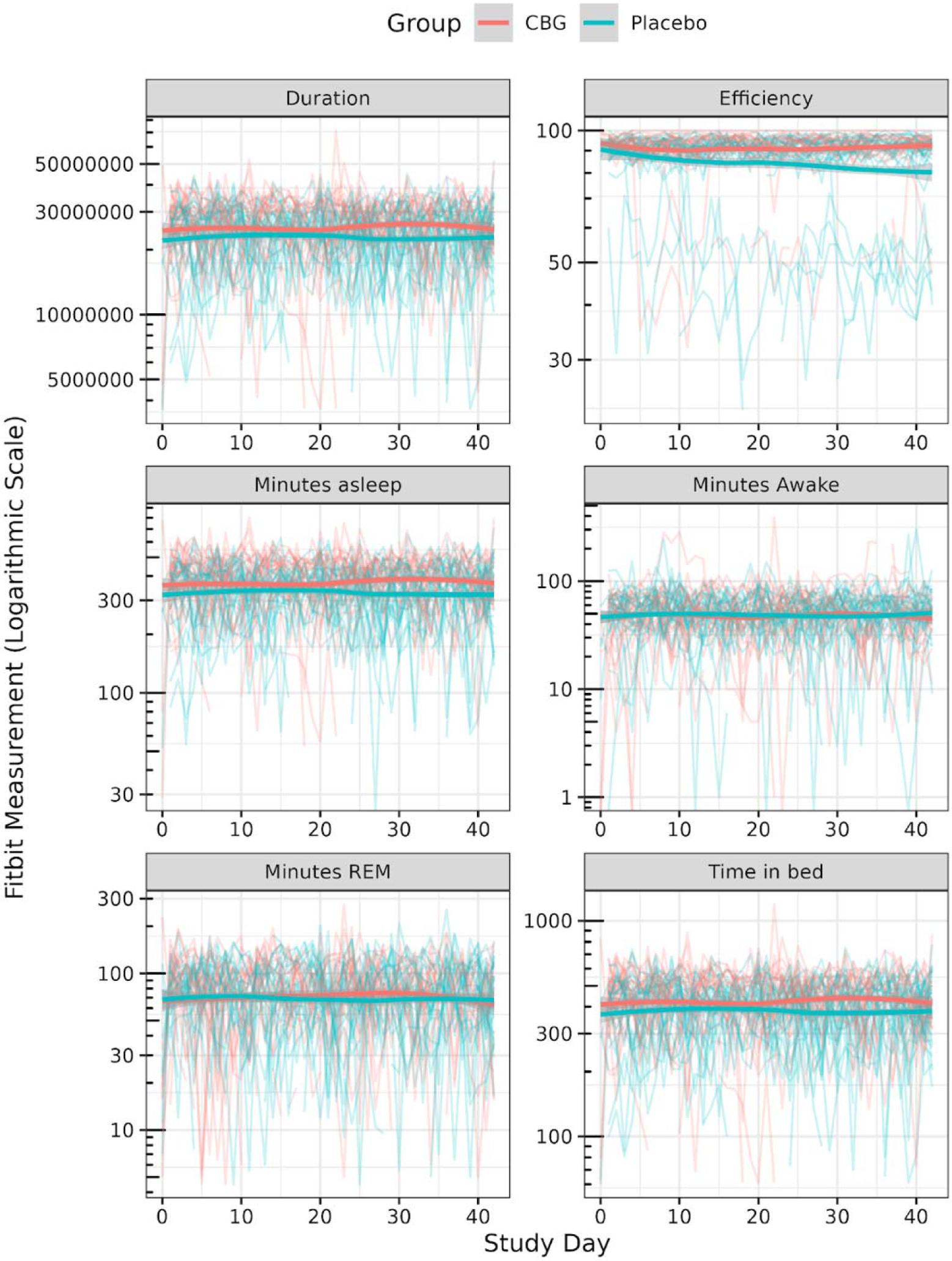
Per-participant daily sleep summaries (per-protocol population) Wrist actigraphy data for sleep duration, efficiency, minutes asleep, minutes awake, minutes of REM, and overall time in bed. Light lines show individual participant summaries, and the bold lines represent smoothed group averages based on loss fit. Abbreviations: CBG, cannabigerol; REM, rapid eye movement

**Figure 6.**
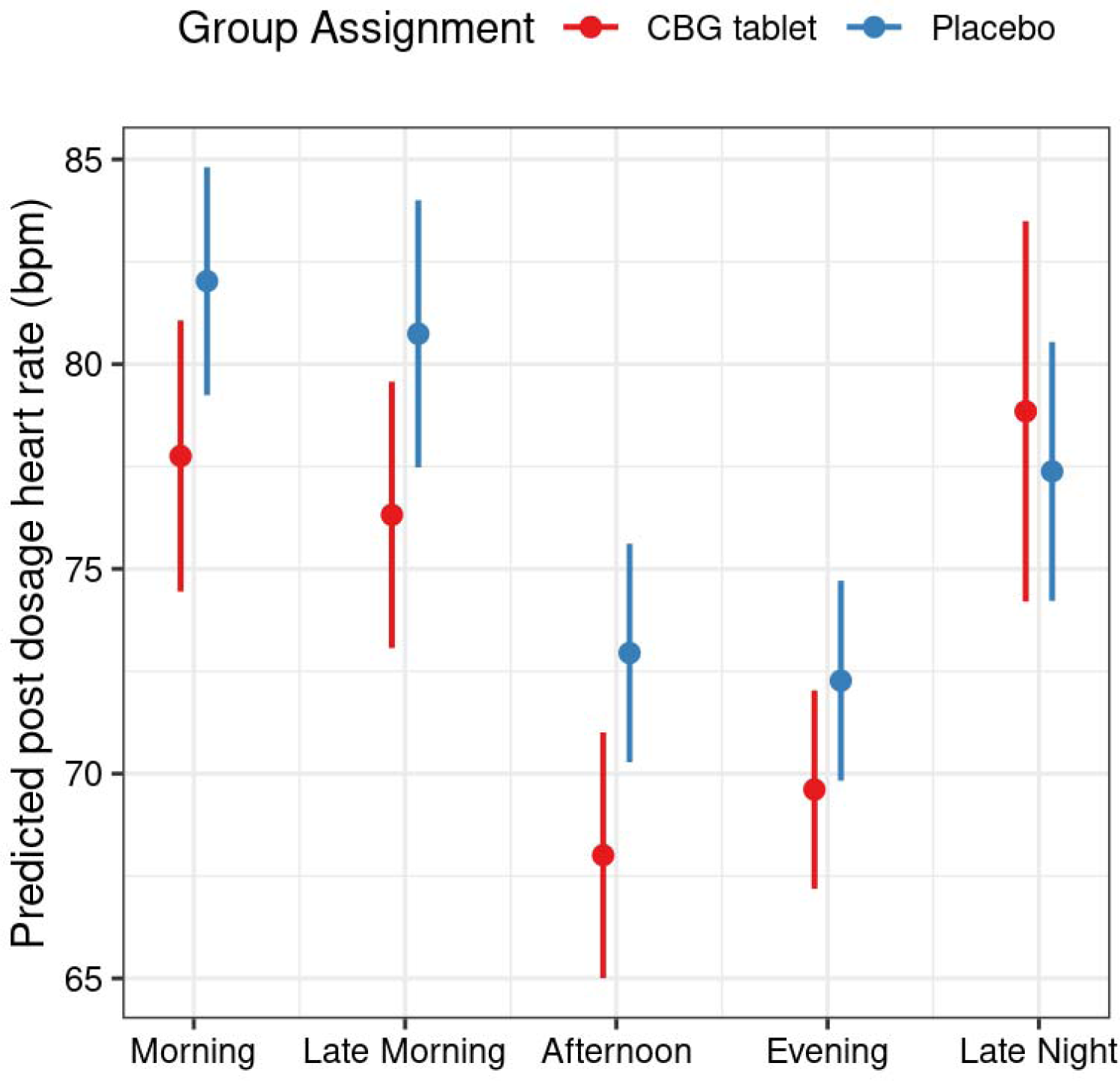
Dosage timing and heart rate (intention-to-treat population) Abbreviations: CBG, cannabigerol

**Table 4.** Physiological effect of CBG dosage timing on resting heart rate (intention-to-treat population)

| Dosing timeframe | Differential (CBG – placebo) |  |  |  |
| --- | --- | --- | --- | --- |
|  | Mean Estimate | Standard Error | 95% Confidence limit | P-value<sup>a</sup> |
| Morning (5 AM – 10 AM) | –4.27 | 2.18 | (–8.58, 0.03) | 0.052 |
| Late morning (10 AM – noon) | –4.42 | 2.33 | (–9.03, 0.18) | 0.060 |
| Afternoon (noon – 5 PM) | –4.94 | 2.05 | (–9.00, –0.88) | 0.017 |
| Evening (5 PM – 9 PM) | –2.66 | 1.75 | (–6.15, 0.83) | 0.133 |
| Late night (9 PM – 5 AM) | 1.47 | 2.85 | (4.15, 7.08) | 0.608 |

<sup>a</sup> p-value for differences between groups calculated using the Wilcoxon rank sum test

### Safety and adverse events

No serious AEs considered possibly related to the study medication were reported. During the study a total of 5 nonserious AEs that were considered possibly related to the study medication were recorded from 5 participants receiving CBG. AEs reported as possibly related to CBG included headache, lethargy, gastric upset, nausea and hypersomnia (one episode for each). All of these AEs were considered mild. While no participant reported using outpatient or emergency medical services for AE management, 2 participants receiving CBG withdrew from further study participation. Two participants (one in each treatment group) reported mild dermatitis/skin irritation related to the Fitbit device.

Emergency alerts triggered by extreme responses to PCL-5 questions addressing negative beliefs or addressing taking risks/self-harm were monitored across the study, with a total of 10 such alerts, involving a total of 9 participants (14%). In those participants receiving CBG, 4 alerts relating to negative beliefs were reported (involving 3 participants) with 2 participants reporting extreme responses relating to taking risks/self-harm. In those receiving placebo, 3 participants reported extreme negative beliefs and one reported an extreme response to the PCL-5 question addressing taking risks/self-harm. After speaking with the on-call clinician, it was determined that none of these participants required emergency or routine medical services, and none were withdrawn from the study.

## Discussion

In this entirely decentralized, randomized, placebo-controlled, triple-blinded trial we evaluated the efficacy and safety of a commercially available CBG formulation in US Veterans with self-reported sleep disturbance. While medicinal cannabis and Δ9-THC and CBD have been studied in a wide range of conditions, including sleep disturbance,^13–19^ to date the clinical data for CBG is sparse, with most studies evaluating effects in-vitro or in animal models.^20^ Although there are some survey data reporting on patterns of CBG use, and adverse effect profile,^23^ to our knowledge this is the first randomized placebo-controlled study evaluating CBG efficacy and safety. In this we found that while use of CBG tended towards improvement in sleep (as evident by decline in MOS-SS SPI-II scores) a similar pattern was observed in the placebo arm, with no statistically discernible difference in scores between the two groups. Similar findings were also observed in the QoL measures although any tendency towards improvements were far smaller. The lower effect on QoL despite improved sleep scores might be explained by other factors (beyond sleep) contributing to broader health issues influencing QoL. Others have reported similar findings in patients using cannabis preparations for problematic sleep.^46^ To this point, PCL-5 scores in the study population were high, and where a high proportion of participants in each group (over 40%) had scores ≥33 (indicative of PTSD) at inclusion. Furthermore, other comorbidities may have influenced QoL and study outcome measures. One limitation of the present study was that, except for OSA, we did not consider medical comorbidities or associated concomitant medications and did not account for these in our randomization or analyses. It was not feasible nor representative to restrict any pharmacological use, including mental health-related or even sleep-related medication.

The two-week run-in phase prior to treatment allowed evaluation of any potential impact of study involvement and participant engagement on outcomes and endpoints. In this context, some improvement in sleep scores was apparent during this pre-treatment period in both groups. This improvement continued across the subsequent four weeks of treatment in both groups, and where decline in MOS-SS SPI-II scores was greater in the placebo group. Substantial placebo responses in randomized studies evaluating cannabinoids for analgesia/pain relief are well recognized, whereby high treatment expectations may impact self-reported outcome measures such as those used in the present study.^47,48^ An additional consideration is the use of cannabis products in our study population (legal within California), with over 75% reporting daily use of Δ9-THC and/or CBD at inclusion. As use could continue throughout the study, this could have masked any additional benefit of CBG. Not restricting cannabis use was a necessary compromise to ensure representative study recruitment and retention in this population.^49^ Future studies in populations without concomitant cannabis product use may allow a clearer understanding of this aspect.

Nevertheless, the present study does provide valuable information. CBG was well tolerated with no serious AEs reported. Five out of 33 participants receiving CBG (15.2%) each experienced one nonserious AE (all considered mild), with two withdrawing from the study. While safety data are limited for CBG, Russo et al. have reported AEs in more than 50% of subjects regularly using CBG products.^23^ In the present study no single AE predominated, with those reported as possibly related to CBG spanning a range of symptoms (headache, lethargy, gastric upset, nausea and hypersomnia), consistent with those previously reported by Russo et al.^23^ This is reassuring, and is supportive of future studies evaluating the CBG formulation. An additional consideration is dosing and treatment duration. Although we did not observe any dose-related response, studies evaluating CBD show greater effects at higher dosing,^50–52^ and it is possible that higher CBG dosing (beyond 50 mg) may achieve better study outcomes. Furthermore, extended dosing beyond 4 weeks could be considered. In the present study, participants could take CBG at any time during the day but no more than three hours before bedtime. It may be that a near-bedtime dosing time may more consistently influence sleep outcomes.

A decentralized study design is increasingly proposed to address inequitable access to clinical trial participation, but poses its own challenge with study retention. A recent cross-study evaluation of 100,000 participants in eight remote digital health studies reported a median participant retention of only 5.5 days.^53^ The present study was entirely decentralized, yet we retained >87% of study participants for the full six-week duration. This affirms the feasibility of randomized controlled trials in underrepresented populations such as Veterans. In addition to the self-reported sleep and QoL outcome measures, we also evaluated passively-collected data via Fitbit on sleep quality, activity tracking and heart rate measures. Such measures are increasingly applied in studies evaluating insomnia, and may provide a useful adjunct to conventional PSG and sleep diaries.^54–56^ Actigraphy data are particularly useful in decentralized studies and merit further comparison to patient-reported outcomes.

## Conclusions

To our knowledge, this is the first completed, randomized placebo-controlled study evaluating efficacy and safety of CBG. While the small sample size and a placebo effect obscured any potential efficacy, CBG was well tolerated. These results support further investigation of the physiological and psychological effects of CBG in future studies.

## Acknowledgements

The authors sincerely appreciate our co-sponsor Veterans Cannabis Group and Jeremy Freitas for their time, effort, and feedback throughout the study design phase. They also thank staff at Curebase Inc. for their role in participant screening and logistical study support. The authors appreciate and acknowledge Irina Angel, M.D. for their role as the independent study clinician and their dedication to study participants. We are particularly grateful for the Veterans that volunteered their time for our participant panels to share their experiences and guide our study design. The authors also thank Iain O’Neill (freelance on behalf of nymbly) for providing medical writing support.

## Data availability statement

The data underlying this article will be shared on reasonable request to the corresponding author.

## Author contributions

C.E.W. and C.R.E. conceptualized and designed the study. E.J.D. developed the Statistical Analysis Plan. B.G.K. performed statistical analysis. C.E.W. wrote the initial manuscript draft. All authors contributed to further manuscript revision and approved the final version.

