## Supplementary for "Effect of cannabigerol on sleep and quality of life in Veterans: A decentralized, randomized, placebo-controlled trial"

^2^ nymbly, Seattle, WA, USA

^3^ Freelance Consultant, Santa Mateo, CA, USA

^4^ Freelance Consultant, c/o nymbly, Seattle, WA, USA

^5^ Freelance Consultant, c/o nymbly, Seattle, WA, USA

***Corresponding author**

Courtney Webster, nymbly, Seattle, WA, USA

**Supplementary Table 1. Inclusion and Exclusion Criteria**

**Inclusion Criteria**

- Veteran status
- MOS Sleep Problems Index II > 30
- California resident
- Age 21 or over
- Participants must own their own device to use for the study. Devices must meet the following criteria:
  - Apple iOS 12.2 or higher
  - Android OS 7.0 or higher
- Participants must be comfortable reading study instructions in English and communicating with study team in English
- Be willing to commit to study dosing, completing evaluation instruments, and following study protocol activities.
- If female and of childbearing potential, agree to use an effective form of birth control during study participation; defined as those which result in a low failure rate (i.e., less than 1% per year) when used consistently and correctly such as implants, injectables, oral contraceptives, IUDs, or a vasectomized partner.
- If using sleep medications, medication and dosage have not been changed in the past month and will remain unchanged for the duration of the study
- If using other psychotropic medications, medication and dosage have not been changed in the past 2 months, and will remain unchanged for the duration of the study.
- If diagnosed with sleep apnea (participant reported), participant must be currently using a CPAP with at least four weeks of prior CPAP use
- If prior observation that the participant has stopped breathing or observed choking/gasping during their sleep, participant must be currently using a CPAP with at least four weeks of prior CPAP use

**Exclusion Criteria**

- Currently in a Cognitive Behavioral Therapy for Insomnia (CBTI) program
- Women who are currently pregnant, trying to become pregnant, or breastfeeding

### **Supplementary Figure 1. Culturally-competent Clinical Research Coordinator Training (Excerpt)**

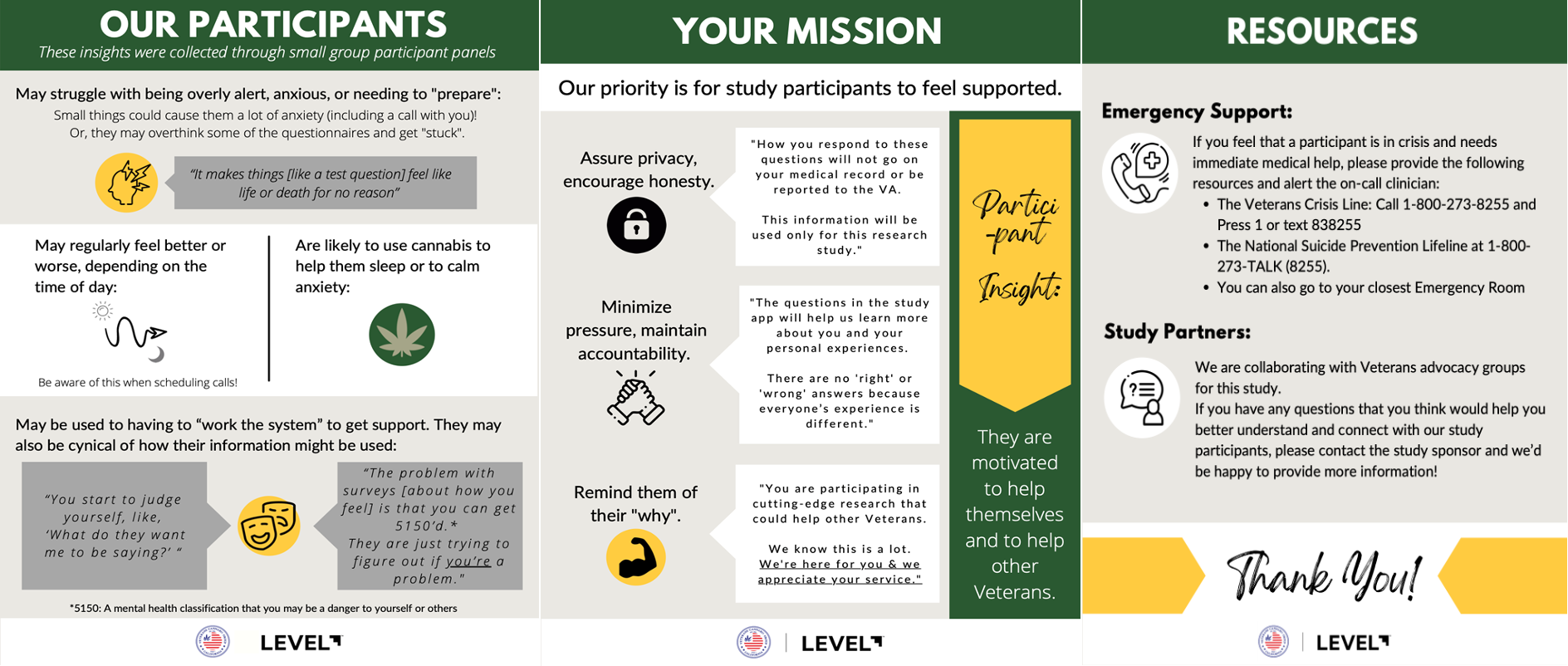

### **Supplementary Figure 2. Schedule of assessments throughout the study**

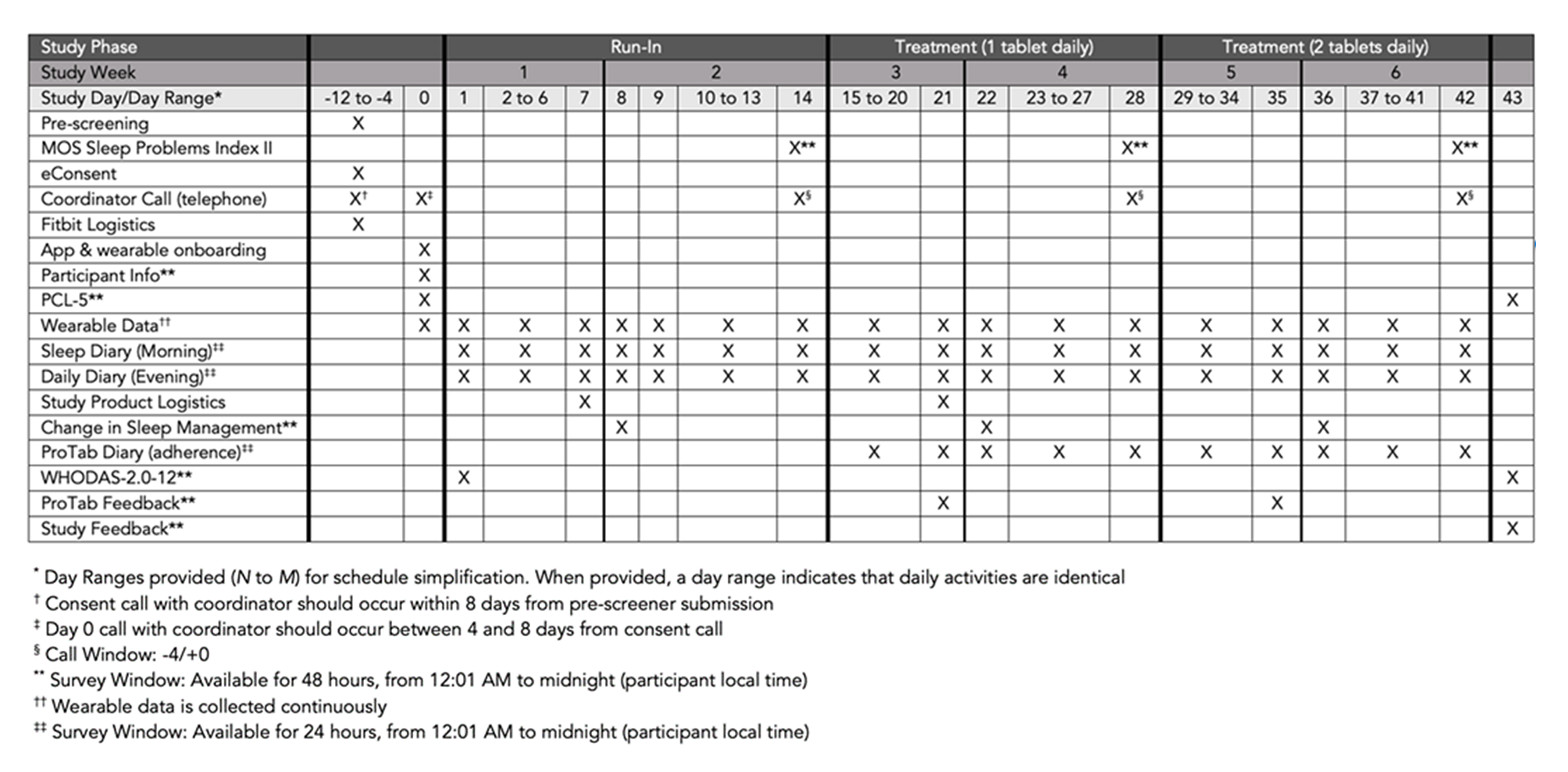
